# Self-reported and physiological reactions to the third BNT162b2 mRNA COVID-19 (booster) vaccine dose

**DOI:** 10.1101/2021.09.15.21263633

**Authors:** Merav Mofaz, Matan Yechezkel, Grace Guan, Margaret L. Brandeau, Tal Patalon, Sivan Gazit, Dan Yamin, Erez Shmueli

## Abstract

**Background:** The rapid rise in hospitalizations associated with the Delta-driven COVID-19 resurgence, and the imminent risk of hospital overcrowding, led the Israeli government to initialize a national third (booster) COVID-19 vaccination campaign in early August 2021, offering the BNT162b2 mRNA vaccine to individuals who received their second dose over five months ago. However, the safety of the third (booster) dose has not been fully established yet.

**Objective:** Evaluate the short-term, self-reported and physiological reactions to the third BNT162b2 mRNA COVID-19 (booster) vaccine dose.

**Design:** A prospective observational study, in which participants are equipped with a smartwatch and fill in a daily questionnaire via a dedicated mobile application for a period of 21 days, starting seven days before the vaccination.

**Setting:** An Israel-wide third (booster) vaccination campaign.

**Participants:** A group of 1,609 (18+ years of age) recipients of at least one dose of the BNT162b2 vaccine between December 20, 2020, and September 15, 2021, out of a larger cohort of 2,912 prospective study participants. 1,344 of the participants were recipients of the third vaccine dose.

**Measurements:** Daily self-reported questionnaires regarding local and systemic reactions, mood level, stress level, sport duration, and sleep quality. Heart rate, heart rate variability and blood oxygen saturation level were continuously measured by Garmin Vivosmart 4 smartwatches.

**Results:** The extent of systemic reactions reported following the third (booster) dose administration is similar to that reported following the second dose (p-value=0.305) and considerably greater than that reported following the first dose (p-value<0.001). Our analyses of self-reported well-being indicators as well as the objective heart rate and heart rate variability measures recorded by the smartwatches further support this finding. Focusing on the third dose, reactions were more apparent in younger participants (p-value<0.01), in women (p-value<0.001), and in participants with no underlying medical conditions (p-value<0.001). Nevertheless, reported reactions and changes in physiological measures returned to their baseline levels within three days from inoculation with the third dose.

**Limitations:** Participants may not adequately represent the vaccinated population in Israel and elsewhere.

**Conclusion:** Our work further supports the safety of a third COVID-19 BNT162b2 mRNA (booster) vaccine dose from both a subjective and an objective perspective, particularly in individuals 65+ years of age and those with underlying medical conditions.

**Primary funding source:** European Research Council (ERC) project #949850

## Introduction

The SARS-CoV-2 Delta variant (also termed variant B.1.617.2) was discovered in October 2020, in India, and was designated as a variant of concern by the World Health Organization (WHO) in May 2021 (1–3). Since its discovery, it has spread worldwide and has rapidly become the most dominant variant in many countries (4–7). Although the BNT162b2 COVID-19 vaccine is highly effective against the Alpha variant (8), recent studies show that the effectiveness of the Pfizer-BioNTech vaccines is notably lower against the Delta variant – 88% compared to 93.7% against the Alpha variant (9–12).

Moreover, recent evidence shows that fully vaccinated individuals infected with the virus can easily transmit it further, as their peak viral burden is similar to that observed in unvaccinated individuals (7,10). In Israel, the Delta variant has accelerated COVID-19 infection and hospitalization, with numbers doubling every ten days between July 1, 2021 and August 9, 2021 (7,13), despite the high coverage of the BNT162b2 vaccine in Israel during this period—greater than 75% coverage with two Pfizer doses in the eligible population (individuals ≥ 12 years of age) (13).

The rapid rise in hospitalizations associated with the Delta-driven COVID-19 resurgence, and the imminent risk of hospital overcrowding, led the Israeli government to initialize on July 30, 2021, an unparalleled, pro-active, national third (booster) vaccine shot campaign, offering the BNT162b2 mRNA COVID-19 vaccine to individuals over the age of 60. On August 13, 2021, the booster campaign was expanded to include those over 50 years of age, reaching 63% third-dose coverage among the eligible population within only 26 days (7,14–16). Two weeks later, on August 29, 2021, the campaign was expanded to include all individuals 16+ of age, demanding only that five months have passed since the receipt of the second dose, reaching 40% third-dose coverage among the eligible population under 50 years of age, within 16 days (13,17).

Currently, limited information is available on the safety of a BNT162b2 third dose (18,19), with such a booster vaccine yet to be authorized by the US Food and Drug Administration (FDA) to the general population (20). While recent evidence shows that a third BNT162b2 dose in immunocompromised individuals has a favorable safety profile (19,21), the safety of a third (booster) dose in the general population has not yet been fully established.

Here, we evaluate the short-term effects of a third BNT162b2 mRNA COVID-19 vaccine dose on self-reported and physiological indicators. We followed a cohort of 2,912 participants; of these, 1,609 participants received at least one dose of the BNT162b2 vaccine after joining the study. Each participant received a dedicated mobile application and a Garmin Vivosmart 4 smartwatch. The mobile application collected daily self-reported questionnaires on local and systemic reactions as well as various well-being indicators. The smartwatch continuously monitored several physiological measures, including heart rate, heart rate variability (HRV) and blood oxygen saturation level. Our analysis of the comprehensive data on each participant examines the safety of a third (booster) vaccine dose from both a subjective perspective (patient questionnaire) and an objective perspective (smartwatch data).

## Materials and Methods

### Study design and participants

Our study includes a prospective cohort of 2,912 participants (18+ years of age) who were recruited between November 1, 2020 and September 15, 2021. The 1,609 participants who reported receipt of at least one of the three BNT162b2 mRNA COVID-19 vaccine shots after joining the study served as the base cohort for our analysis. Specifically, of these 1,609 participants, during the study, 223 received their first dose, 351 their second dose, and 1,344 their third dose.

We employed a professional survey company to recruit participants and ensure they follow through with the study requirements. Participant recruitment was performed via advertisements on social media and word-of-mouth. Each participant provided informed consent by signing a form after receiving a comprehensive explanation on the study. Then, participants completed a one-time enrollment questionnaire, were equipped with Garmin Vivosmart 4 smartwatches, and installed two applications on their mobile phones: (1) the PerMed application (22) that collected daily self-reported questionnaires, and (2) an application that passively recorded the smartwatch data. Participants were asked to wear their smartwatches as much as possible. The survey company ensured that participants’ questionnaires were completed daily, that their smartwatches were charged and properly worn, and that any technical problems with the mobile applications or smartwatch were resolved.

### PerMed mobile application

Participants used the PerMed mobile application (22) to fill out daily questionnaires. The questionnaire allowed participants to report various well-being indicators, including mood level (on a scale of 1 [awful] to 5 [excellent]), stress level (on a scale of 1 [very low] to 5 [very high]), sport duration (in minutes), and sleep quality (on a scale of 1 [awful] to 5 [excellent]). The questionnaire also collected data on clinical symptoms consistent with the local and systemic reactions observed in the BNT162b2 mRNA Covid-19 clinical trial (23), with an option to add other symptoms as free text. (For more details see Appendix A in the Supplemental Material)

We implemented preventive measures to improve the quality, continuity, and reliability of the collected data. Each day, if by 7 pm, participants had not yet filled out the daily questionnaire, they received a reminder notification through the PerMed application. Furthermore, we used a dedicated dashboard to identify participants who continually neglected to complete the daily questionnaires; these participants were contacted by the survey company and were encouraged to better adhere to the study protocol.

### The smartwatch

Participants were equipped with Garmin Vivosmart 4 smart fitness trackers. Among other features, the smartwatch provides all-day heart rate and heart rate variability and during-night blood oxygen saturation level tracking capabilities (24).

The optical wrist heart rate (HR) monitor of the smartwatch is designed to continuously monitor a user’s heart rate. The frequency at which heart rate is measured varies and may depend on the level of activity of the user: when the user starts an activity, the optical HR monitor’s measurement frequency increases.

Since heart rate variability (HRV) is not easily accessible through Garmin’s application programming interface (API), we use Garmin’s stress level instead, which is calculated based on HRV. Specifically, the device uses heart rate data to determine the interval between each heartbeat. The variable length of time between each heartbeat is regulated by the body’s autonomic nervous system. Less variability between beats correlates with higher stress levels, whereas an increase in variability indicates less stress (25). A similar relationship between HRV and stress was also seen in (26,27).

The Pulse Ox monitor of the smartwatch uses a combination of red and infrared lights with sensors on the back of the device to estimate the percentage of oxygenated blood (peripheral oxygen saturation, SpO2%). The Pulse Ox monitor is activated each day at a fixed time for a period of four hours (the default is 2AM-6AM).

Examining the data collected in our study, we identified an HR sample roughly every 15 seconds, an HRV sample every 180 seconds, and an SpO2 sample every 60 seconds.

While the Garmin smartwatch provides state-of-the-art wrist monitoring, it is not a medical-grade device, and some readings may be inaccurate under certain circumstances, depending on factors such as the fit of the device and the type and intensity of the activity undertaken by a participant (28–30).

### Statistical analysis

Questionnaire data were preprocessed by manually categorizing any self-reported symptom entered as free text. In addition, if participants filled out the questionnaire more than once in one day, the last entry from that day was used in the analysis. Smartwatch data were preprocessed as follows. First, we computed the mean value of each hour of data. We then performed linear interpolation to impute missing hourly means. Lastly, we smoothed the data by calculating the five-hour moving average.

For each participant, we defined the 7-day period prior to vaccination as the baseline period. We noted any pre-existing clinical symptoms from the last questionnaire completed during the baseline period. Next, we calculated the percentage of participants who reported new (that is, not pre-existing) systemic reactions in the 48 hours after vaccination. For each reaction, we used a beta distribution to determine a 90% confidence interval. To determine the statistical significance of differences between the first and third doses and between the second and third doses as reflected by the extent of reported reactions, a two proportion Z-test was used.

We also calculated the mean difference in well-being indicators between the post-vaccination period and the baseline period. Specifically, for each indicator, for each of the three days post-vaccination, and for each participant, we calculated the difference between that indicator’s value and its corresponding value in the baseline period. Then, we calculated the mean value over all participants and the associated 90% confidence interval.

To compare the changes in smartwatch physiological indicators over the seven days (168 hours) post vaccination with those of the baseline period, we performed the following steps. First, for each participant and each hour during the seven days post vaccination, we calculated the difference between that hour’s indicator value and that of the corresponding hour in the baseline period (keeping the same day of the week and same hour during the day). Then, we aggregated each hour’s differences over all participants to calculate a mean difference and associated 90% confidence interval, which is analogous to a one-sided t-test with significance level 0.05. To determine the statistical significance of differences between the first and third doses and between the second and third doses as reflected by changes in smartwatch indicators during the 48 hours post-vaccination, we used a two-sample t-test with unequal variance.

We repeated the above analyses for the third dose stratified by age groups (<50, 50-64, and ≥65), gender and the existence of an underlying medical condition (present vs. not present) from the list specified in Table 1.

**Table 1.** Characteristics of the participants

| Characteristic | All Participants<br>(N=1,609) | First Dose<br>(N=223) | Second Dose<br>(N=351) | Third Dose<br>(N=1,344) |
| --- | --- | --- | --- | --- |
| **Gender** |  |  |  |  |
| Male | 46.92% (755) | 45.29%<br>(101) | 45.58% (160) | 47.54% (639) |
| Female | 53.08% (854) | 54.71%<br>(122) | 54.42% (191) | 52.46% (705) |
| **Age group** |  |  |  |  |
| 18-29 yr | 14.23% (229) | 6.28% (14) | 11.11% (39) | 14.06% (189) |
| 30-39 yr | 16.9% (272) | 4.93% (11) | 15.1% (53) | 16.29% (219) |
| 40-49 yr | 11.0% (177) | 6.73% (15) | 11.97% (42) | 10.27% (138) |
| 50-59 yr | 26.1% (420) | 28.7% (64) | 24.79% (87) | 27.9% (375) |
| 60-69 yr | 22.25% (358) | 31.39% (70) | 21.37% (75) | 22.92% (308) |
| ≥ 70 yr | 9.51% (153) | 21.97% (49) | 15.67% (55) | 8.56% (115) |
| **Body-mass index\*** |  |  |  |  |
| < 30.0 | 78.19% (1,258) | 78.48%<br>(175) | 79.77% (280) | 77.68% (1,044) |
| ≥ 30.0 | 20.51% (330) | 18.39% (41) | 17.09% (60) | 21.43% (288) |
| Unspecified | 1.31% (21) | 3.14% (7) | 3.13% (11) | 0.89% (12) |
| **Underlying Medical Condition** |  |  |  |  |
| Hypertension | 14.17% (228) | 20.63% (46) | 15.95% (56) | 14.43% (194) |
| Diabetes | 8.64% (139) | 13% (29) | 7.98% (28) | 8.41% (113) |
| Heart disease | 4.79% (77) | 7.17% (16) | 4.56% (16) | 4.99% (67) |
| Chronic lung disease | 5.03% (81) | 4.93% (11) | 3.7% (13) | 5.21% (70) |
| Immune suppression | 0.81% (13) | 1.35% (3) | 0.85% (3) | 0.89% (12) |
| Cancer | 0.62% (10) | 0.45% (1) | 0.57% (2) | 0.67% (9) |
| Renal failure | 0.5% (8) | 1.79% (4) | 1.42% (5) | 0.45% (6) |
| None of the above | 73.34% (1,180) | 64.57%<br>(144) | 72.08% (253) | 73.21% (984) |
| Unspecified | 1.06% (17) | 1.35% (3) | 2.85% (10) | 0.52% (7) |

\* The body-mass index is the weight in kilograms divided by the square of the height in meters

### Ethical Approval

Before participating in the study, all subjects were advised, both orally and in writing, as to the nature of the study and gave written informed consent. The study was approved by MHS’ Helsinki institutional review board, protocol number 0122-20-MHS.

## Results

Of the 1,609 participants who received at least one dose of the BNT162b2 vaccine after joining the study, 854 (53.08%) were women, and 755 (46.92%) were men. Their age ranged between 18 and 88 years, with a median age of 52 (Table 1). 1,258 (78.19%) participants had a body mass index below 30, and 412 (25.61%) had at least one specific underlying medical condition (Table 1). The distributions of age and gender and the existence of underlying medical conditions were relatively invariable across the recipients of the first, second and third dose (Table 1).

Our examination of self-reported reactions revealed that the extent of systemic reactions reported following the third vaccine dose are similar to those reported following the second dose (p-value=0.305), and considerably greater than those observed following the first dose (p-value<0.001) (Figure 1). Specifically, 60.4% (90% CI: 57.9-62.9% (of the participants did not report any new symptoms after receiving the third dose, compared to 86.5% (90% CI: 81.9-91.0% (and 63.6% (90% CI: 59.1-67.8% (after the first and second doses, respectively. Moreover, the most frequently reported types of reactions—fatigue, headache, muscle pain, fever, and chills—were similar after the second and third doses. These reactions faded in nearly all participants within three days (Figure S8 in the Supplemental Material). Notably, these trends are consistent with those reported in the first and second dose BNT162b2 mRNA vaccine clinical trial (23).

**Figure 1.**
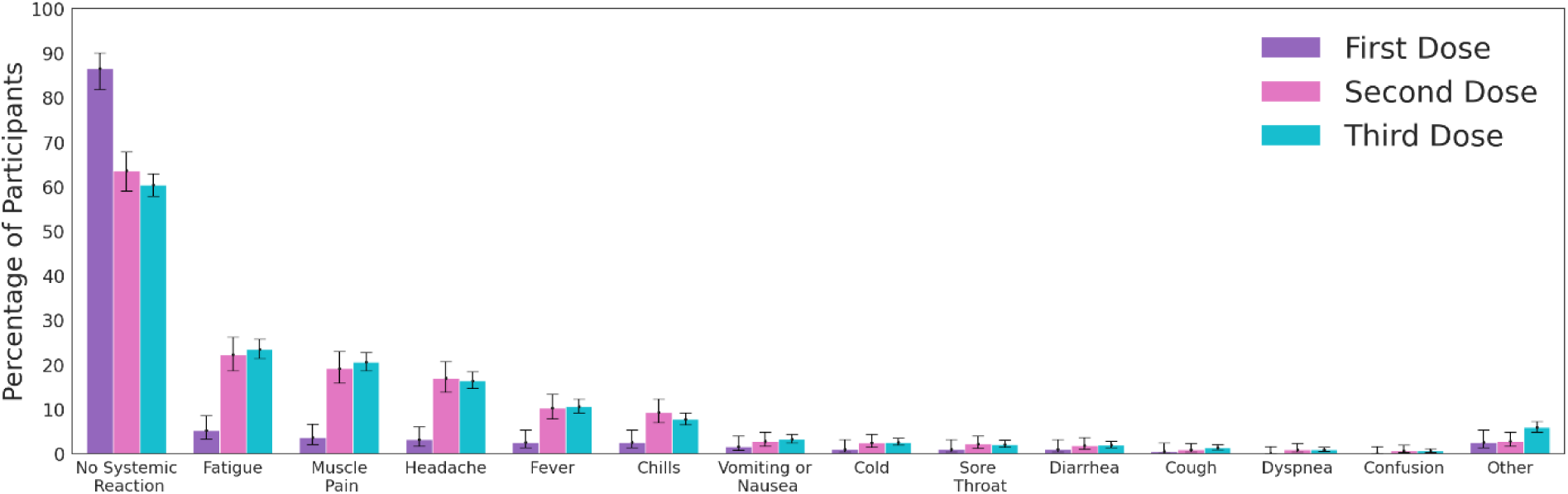
Reactions reported by participants through the mobile application. The bars represent the percentage of participants who reported a given symptom. Error bars represent 90% confidence intervals.

For the self-reported well-being indicators (Fig. 2), we found that during the first two days after the third vaccine dose, participants exhibited a significant reduction in mood level (Fig. 2A), sport duration (Fig. 2C), sleep quality (Fig. 2D) and a notable increase in stress level (Fig. 2B) compared to baseline levels. These changes faded away on the third day post vaccination. A similar trend can be observed after the second vaccine dose, except for the reported stress level that remains below the baseline level during the second and third days post-vaccination.

**Figure 2.**
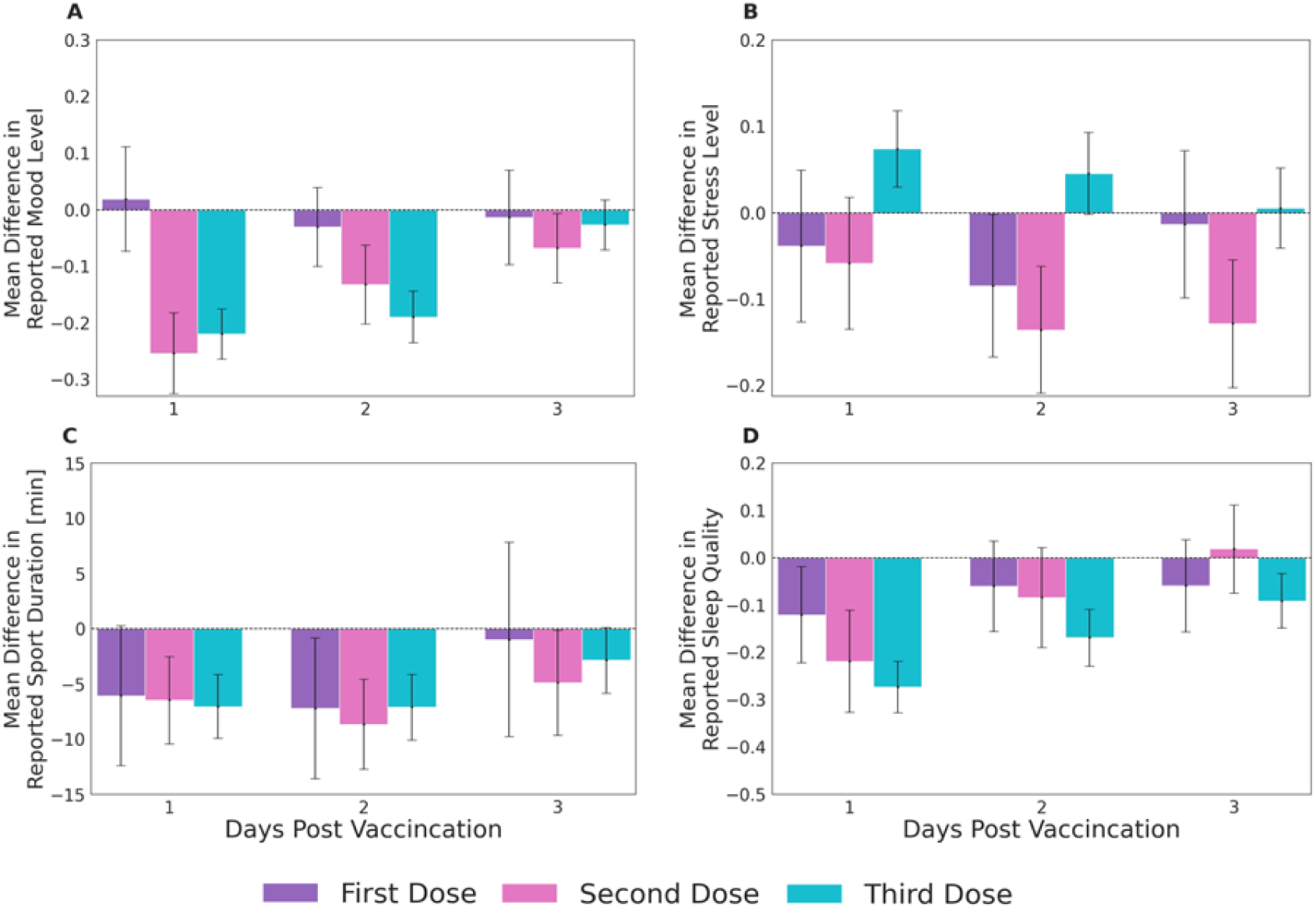
Changes in subjective well-being indicators reported by participants through the mobile application. Mean difference compared to baseline levels for the well-being indicators of (A) Mood level, (B) Stress level, (C) Sport duration, and (D) Sleep quality. Mood level, Stress level, and Sleep quality were reported on a 1-5 Likert scale. Sport duration was measured in minutes. Error bars represent 90% confidence intervals. Horizontal dashed lines represent no change compared to baseline levels.

We observed similar trends when analyzing the objective and continuous physiological measurements collected by the smartwatch (Fig. 3 below and Fig. S1 in the Supplemental Material). Specifically, we identified a considerable rise in both the heart rate (Fig. 3A-C) and the heart rate variability-based stress (Fig. 3D-F) indicators in the first 48 hours following the administration of the third dose. Measurements returned to their baseline levels within 72 hours. By contrast, our analysis of blood oxygen saturation levels suggests that there are no apparent changes following inoculation compared to baseline levels (Fig. 3G-I). The trends observed for the objective heart rate (HR) and heart rate variability (HRV) indicators were consistent with those of the subjective indicators in the following sense - similar changes following the second and third doses (HR p-value=0.43, HRV p-value=0.28), and greater changes following the second and third doses than the first dose (HR p-value<0.03, HRV p-value<0.01).

**Figure 3.**
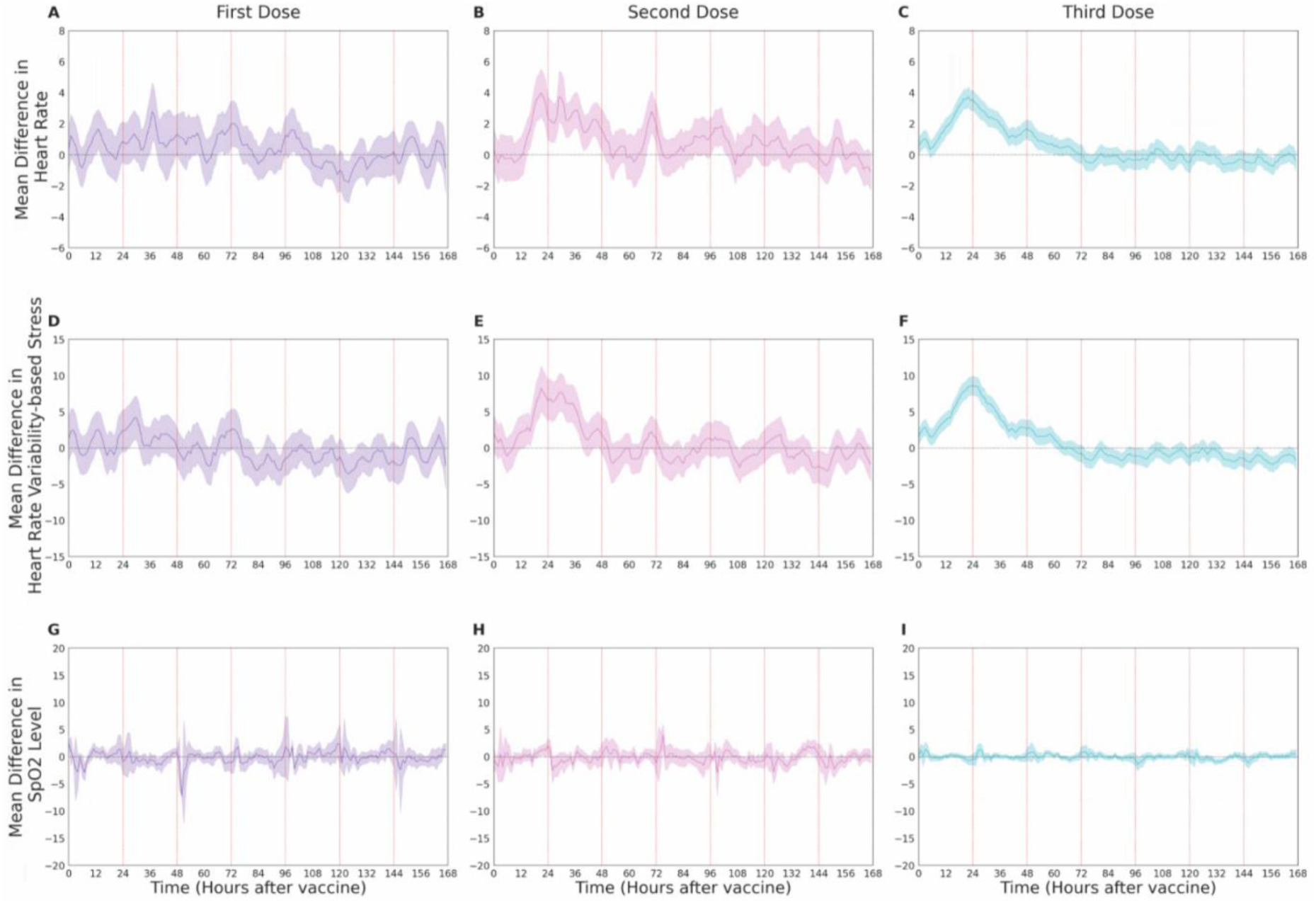
Changes in objective physiological indicators measured through the smartwatch. Mean difference in smartwatch-recorded (**A-C**) heart rate, (**D-F**) heart rate variability-based stress and (**G-I**) blood oxygen saturation level (SpO2) following the first, second and third dose, compared to their baseline levels. Mean values are depicted as solid lines; 90% confidence intervals are presented as shaded regions. The horizontal dashed line represents no change compared to the baseline levels, and vertical lines represent 24-hour periods.

We also stratified our analyses of well-being and smartwatch physiological indicators after the third vaccination by age group, gender, and the existence of a prior underlying medical condition (Fig. 4 below and Fig. S2-S7 in the Supplemental Material). For all stratifications, trends were similar to those observed in the general population. Namely, there were considerable changes in the two days after vaccine administration that faded almost entirely after three days. We also found that participants 65+ years reported fewer reactions (p-value<0.001) and exhibited milder physiological changes compared to those between 50-65 years (HR p-value=0.02, HRV p-value=0.07), and participants between 50-65 years reported even fewer reactions (p-value<0.01) and exhibited milder physiological changes in heart rate (HR p-value=0.02), compared to those below 50 years (Figure 4A-B). Male participants reported fewer reactions (p-value<0.001) but did not exhibit milder physiological changes (HR p-value=0.37, HRV p-value=0.59) than female participants. Participants with an underlying medical condition reported fewer reactions (p-value<0.001) and exhibited milder physiological changes (HR p-value=0.04, HRV p-value=0.16), compared to those without an underlying medical condition. Markedly, out of 9 participants who reported dyspnea 4 (0.96% of their age group) were below 50 years, 4 (0.93% of their age group) were between 50-64 years and 1 (0.65% of her age group) was 65+ years. One participant below 50 years reported chest pain following inoculation. None of these participants had underlying medical condition. These reactions (i.e., dyspnea and chest pain) disappeared between 2-4 days following inoculation.

**Figure 4.**
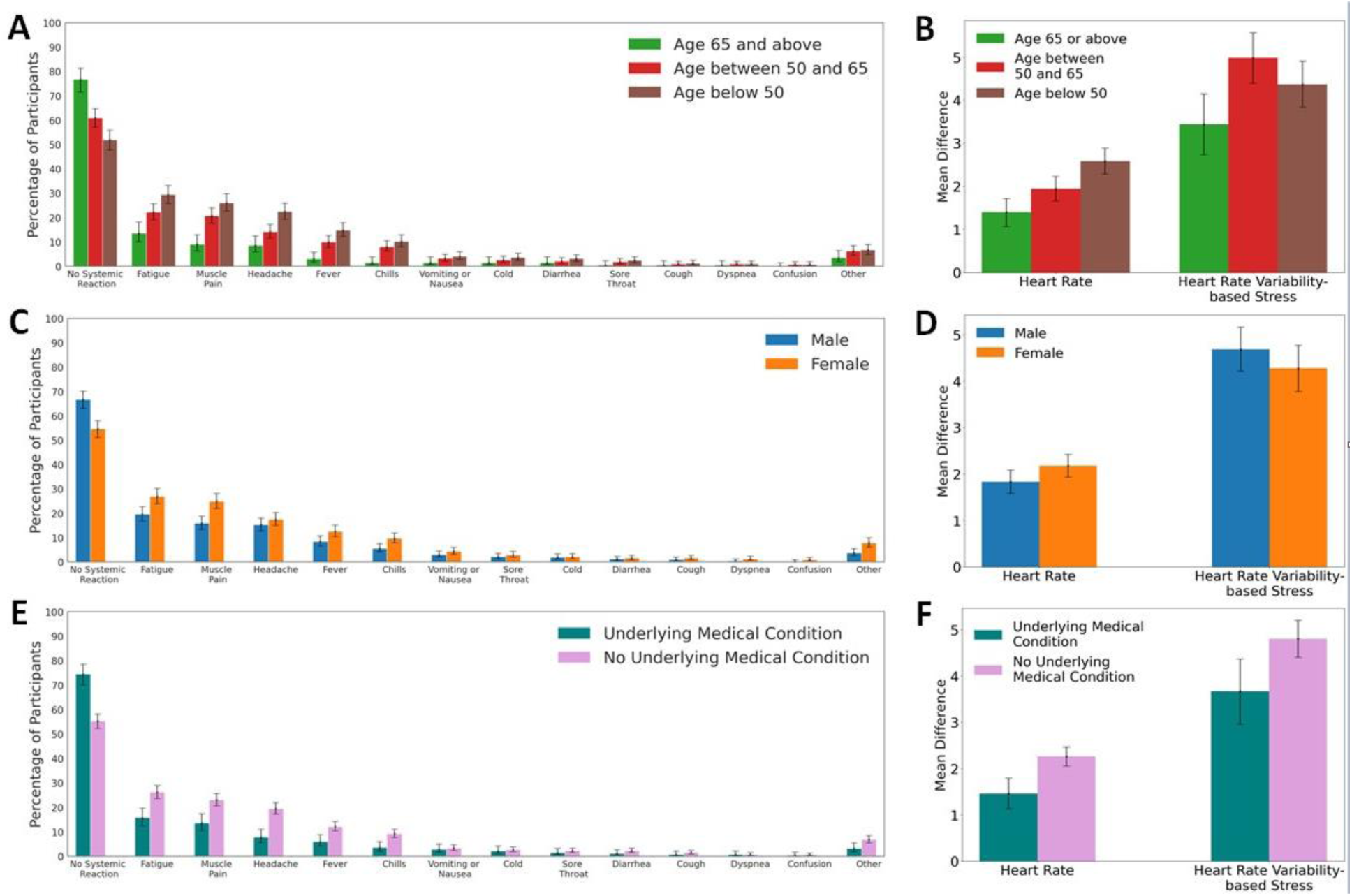
Self-reported and objective reactions following the third vaccine dose, stratified by age, gender and the existence of an underlying medical condition. Reactions reported by participants through the mobile application, stratified by age (A), gender (B) and underlying medical condition (E). The bars represent the percentage of participants who reported a given symptom. Error bars represent 90% confidence intervals. Changes in objective heart rate and heart rate variability measured through the smartwatch in the 48 hours post-vaccination, stratified by age (B), gender (D) and underlying medical condition (F).

## Discussion

Our key findings suggest that local and systemic reactions reported following the third (booster) vaccine dose administration are very similar to those reported following the second dose and considerably greater than those observed following the first dose. Our analyses of both self-reported well-being indicators and objective smartwatch physiological indicators underscore these results. Furthermore, within three days from inoculation with the third dose, all measures returned to their baseline levels in nearly all participants.

We identified differences in subpopulations based on gender, age and underlying medical condition following administration of the third vaccine dose. It has been previously suggested that reactions caused by the COVID-19 vaccine are a byproduct of a short burst of IFN-I generation concomitant with induction of an effective immune response (31). IFN-I generation is substantially stronger in females than in males and stronger in younger and healthier individuals than in older and less healthy ones. We found that participants below the age of 65, females, and those without an underlying medical condition experienced greater reactions both in self-reported local and systemic reactions and well-being indicators as well as in objective physiological measurements recorded by the smartwatch. Our results are also in line with a previous study that found similar trends after the first and second doses (32).

Clinical trials have not yet capitalized on the comprehensive physiological measures generated by wearables. Currently, the FDA and European Medicines Agency (EMA) evaluate the safety and create guidelines for newly developed vaccines primarily based on subjective, self-reported questionnaires (33,34). Much of the scientific literature discusses these self-reported side effects of COVID-19 vaccines. However, integrating wearables into clinical trials, alongside self-reported questionnaires, can provide more precise and rich data regarding the vaccines’ effects on physiological measures.

Our study has several limitations. First, the 1,609 individuals who comprise the base cohort of our analyses may not be representative of the vaccinated population in Israel or globally. Nevertheless, the changes observed in self-reported reactions and well-being indicators as well as objective physiological indicators recorded by the smartwatches were statistically significant and consistent with each other. Moreover, the reaction types, frequency and duration we observed for the first and second dose were similar to those observed in the BNT162b2 mRNA vaccine clinical trials (23). In addition, a clear pattern of returning to baseline levels was observed within 72 hours post inoculation in all examined measures. Lastly, although the sample size was limited, trends were consistent regardless of age group, gender, and the existence of underlying medical conditions.

Second, we did not explicitly control for the effects of the observational trial setting (i.e., participating in a trial, wearing a smartwatch, potential concerns regarding the vaccine, etc.). Any effects of the observational trial setting should, in principle, have similar impacts on our analysis of each of the three vaccine doses. However, since we found no deviations in most measurements from baseline levels in the subset of participants who received their first dose, we believe the changes observed after the second and third doses arise from an actual reaction to the vaccine.

Third, the smartwatches used to obtain physiological measurements are not medical-grade devices. Nevertheless, recent studies show a considerably accurate heart rate measurement in the previous versions of the smartwatch used in this study (28,29). Moreover, our analysis focused on the change in measurements compared to their baseline values rather than on their absolute values.

Finally, the only vaccine available in Israel is the BNT162b2 mRNA vaccine. Although our findings may not directly generalize to other types of COVID-19 vaccines, we believe that applying our analyses on other vaccines are likely to yield qualitatively similar findings, due to the similarities observed between different COVID-19 vaccines (23,35,36).

Our study strengthens the evidence regarding the short-term safety of the booster BNT162b2 vaccine in several ways. First, reports of local and systemic reactions following the third dose were very similar to those observed following the second dose, which was already shown in clinical trials to be safe (23). Second, the considerable changes observed in all indicators during the first two days after receiving the third vaccine, including self-reported reactions and well-being indicators as well as objective physiological indicators collected by the smartwatch, returned to their baseline levels. Third, regardless of the observed differences between subpopulations, our analyses indicate a clear pattern of return to baseline levels in all considered subpopulations. Fourth, we observed no change in blood oxygen saturation levels compared to baseline levels, indicating that major adverse health consequences are less likely.

In conclusion, our study supports the short-term safety of the third BNT162b2 mRNA COVID-19 (booster) vaccine dose and mitigates, in part, concerns regarding its short-term effects. The medical and scientific communities could greatly benefit from the largely unbiased data generated by digital health technologies such as the wearable data which we analyzed in this study. Our findings could also be of interest to public health officials and other stakeholders, as it is important that objective measures are given attention in the critical evaluation of clinical trials.

## Data Availability

Researchers interested in obtaining an aggregated version of the data sufficient to reproduce the results reported in this paper should contact the corresponding author.

## Financial Support

This research was supported by the European Research Council (ERC) project #949850 and the Israel Science Foundation (ISF), grant No. 3409/19, within the Israel Precision Medicine Partnership program. MLB and GG received support from the Koret foundation grant for

Smart Cities and Digital Living 2030.

## Authors’ contributions

Conception and design: DY, ES. Collection and assembly of data: MM, TP, SG, ES. Analysis and interpretation of the data: MM, GG, MLB, DY, ES. Statistical expertise: MM, DY, ES. Drafting the article: MM, MY, DY, ES. Critical revision of the article for important intellectual content: DY, ES. Final approval of the article: All authors. Obtaining funding: DY, ES.

## Reproducible Research Statement

Further details are available from Dr. Erez Shmueli (e-mail). Statistical code will be available after acceptance.

## Supplementary Materials

## Appendix A: Self-reported questionnairs

### Enrollment questionnaire

All participants will fill a one-time enrollment questionnaire that includes demographic questions and questions about the participant’s health condition in general. Specifically, the questionnaire will include the following: age, gender, height, weight and underlying medical conditions (Listed in Table 1, main text). Other questions such as name, address, phone number and email will be recorded and used by the survey company to contact the participants. The answers will be filled-in directly by the survey company to the study’s secured dashboard.

### Vaccination questionnaire

The vaccination questionnaire we will use includes the following question:

- COVID-19 vaccination – date, time and dose number.

### Daily questionnaire

All participants will complete the daily self-reported questionnaire in a dedicated application (the PerMed mobile application). The daily questionnaire we will use includes the following questions:

- How is your mood today? • Awful (1) • Bad (2) • OK (3) • Good (4) • Excellent (5)
- How would you describe the level of your stress during the last day? • Very Low (1) • Low (2) • Medium (3) • High (4) • Very high (5)
- How would you define your last night sleep quality? • Awful (1) • Bad (2) • OK (3) • Good (4) • Excellent (5)
- Try to remember how many minutes of sports activity you performed on the last day?
- Have you experienced one or more of the following symptoms in the last 24 hours? • My general feeling is good, and I have no symptoms • Heat measured above 37.5 • Cough • Sore throat • Runny nose • Headache • Shortness of breath • Muscle aches • Weakness / fatigue • Diarrhea • Nausea / vomiting • Chills • Confusion • Loss of sense of taste / smell • Other symptom.

## Appendix B: Additional Results

### Changes in objective physiological indicators measured through the smartwatch following the three vaccine doses

Changes in objective physiological indicators observed during the first two days after the second and third vaccine doses are similar, and considerably greater than those observed following the first dose.

**Figure S1.**
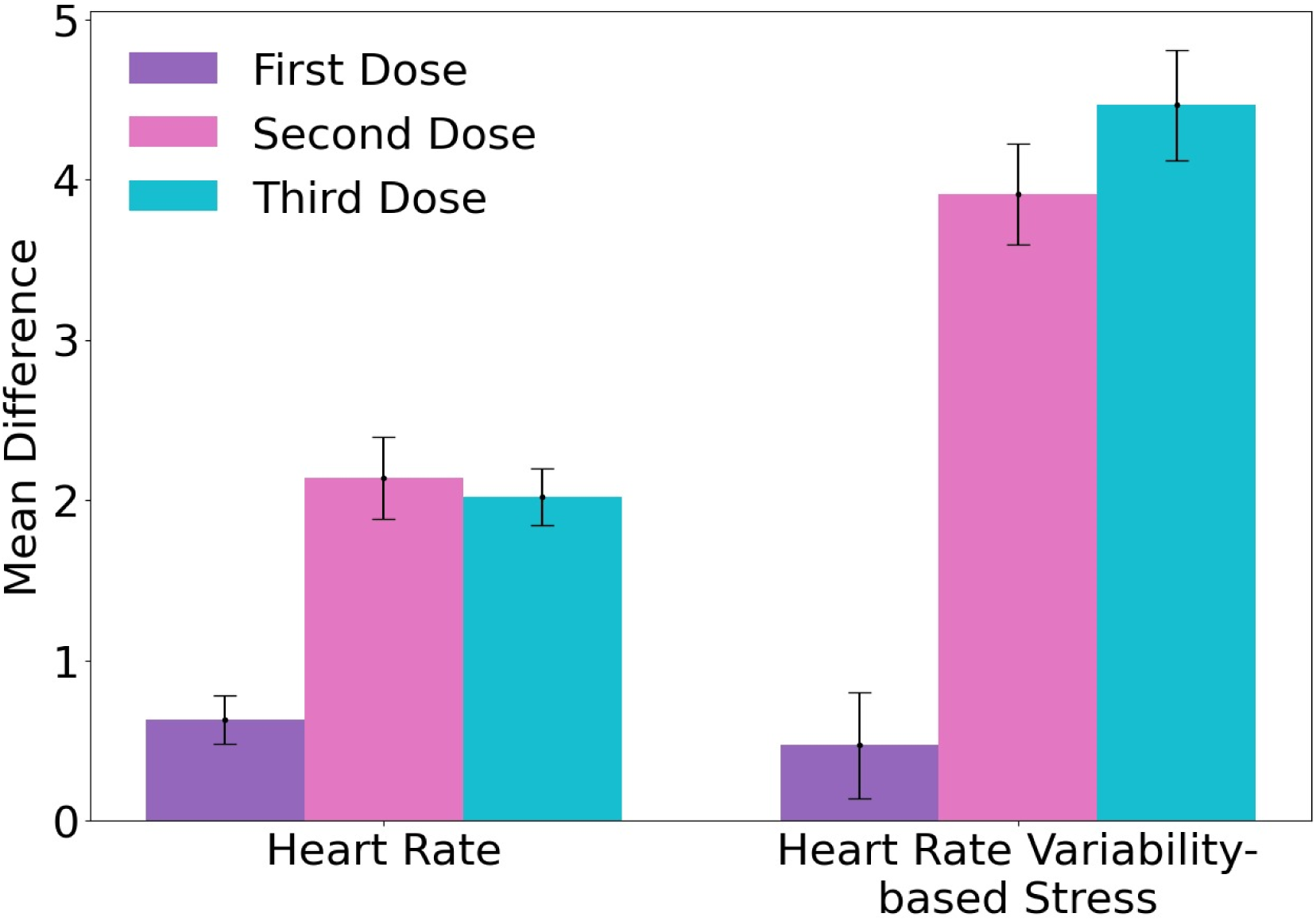
Changes in objective physiological indicators measured through the smartwatch during the first two days after vaccine. Mean difference in smartwatch-recorded heart rate and heart rate variability-based following the first, second and third dose, compared to their baseline levels. Changes in objective physiological indicators were calculated by subtracting the baseline values from the mean value of the first two days following the vaccine dose. Error bars represent 90% confidence intervals.

### Changes in reported well-being indicators – stratification by age group

Changes in well-being observed during the first two days after the third vaccine dose were found to be higher for participants younger than 50 years compared to those between 50 and 65 years, and consequently higher than those older than 65 years, with the exception of reported stress level (Figure S2).

**Figure S2.**
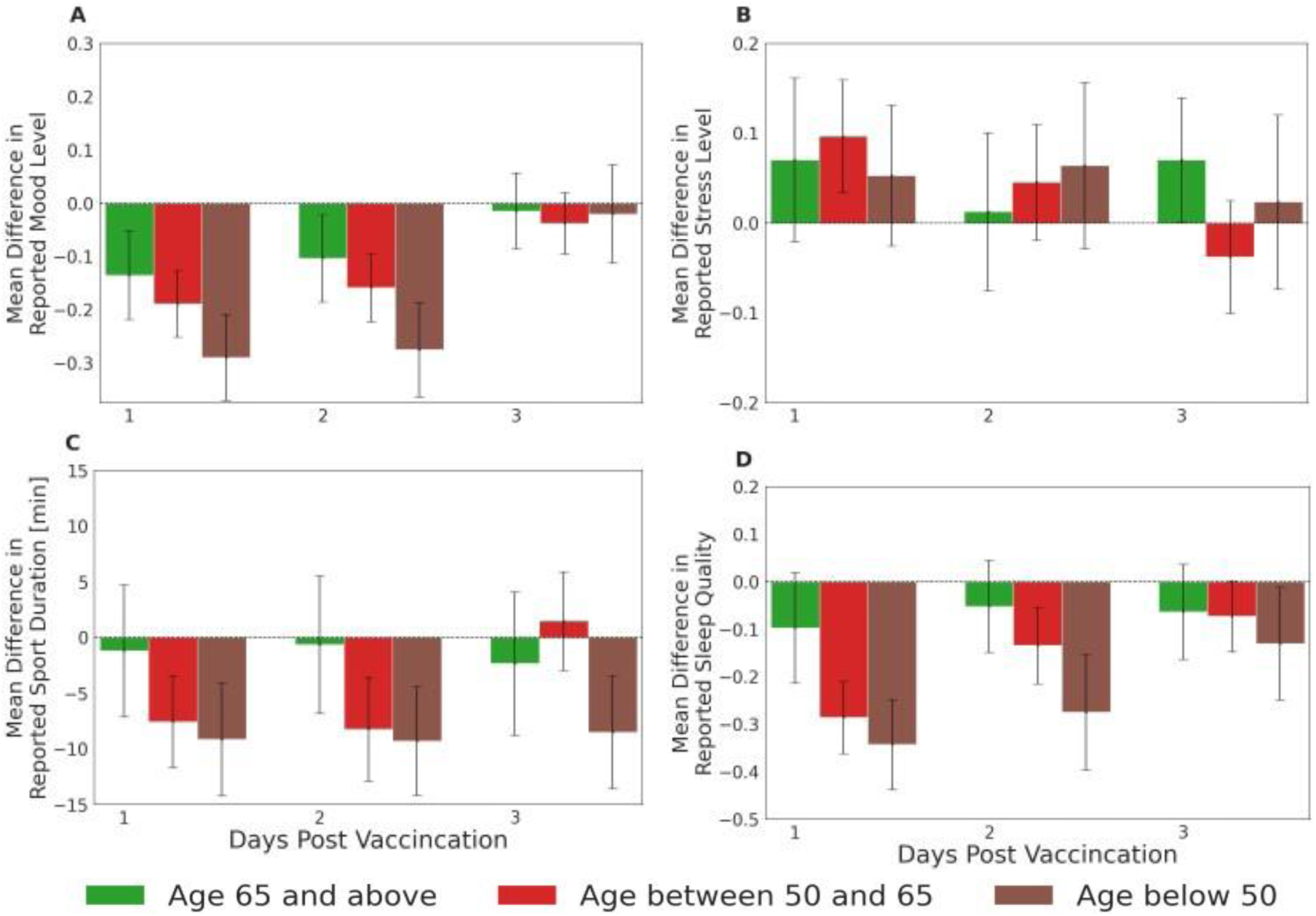
Changes in well-being indicators reported by participants through the mobile application stratified by age group. (A) mood level, measured on a 1-to-5 Likert scale. (B) Stress level, measured on a 1-to-5 Likert scale. (C) Sport duration, measured in minutes. (D) Sleep quality, measured on a 1-to-5 Likert scale. Changes in well-being indicators were calculated by subtracting the baseline values from the daily values. Error bars represent 90% confidence intervals. Horizontal dashed lines represent no change compared to baseline levels.

### Changes in reported well-being indicators – stratification by gender

Changes in well-being observed during the first two days after the third vaccine dose were found to be similar for males and females (Figure S3).

**Figure S3.**
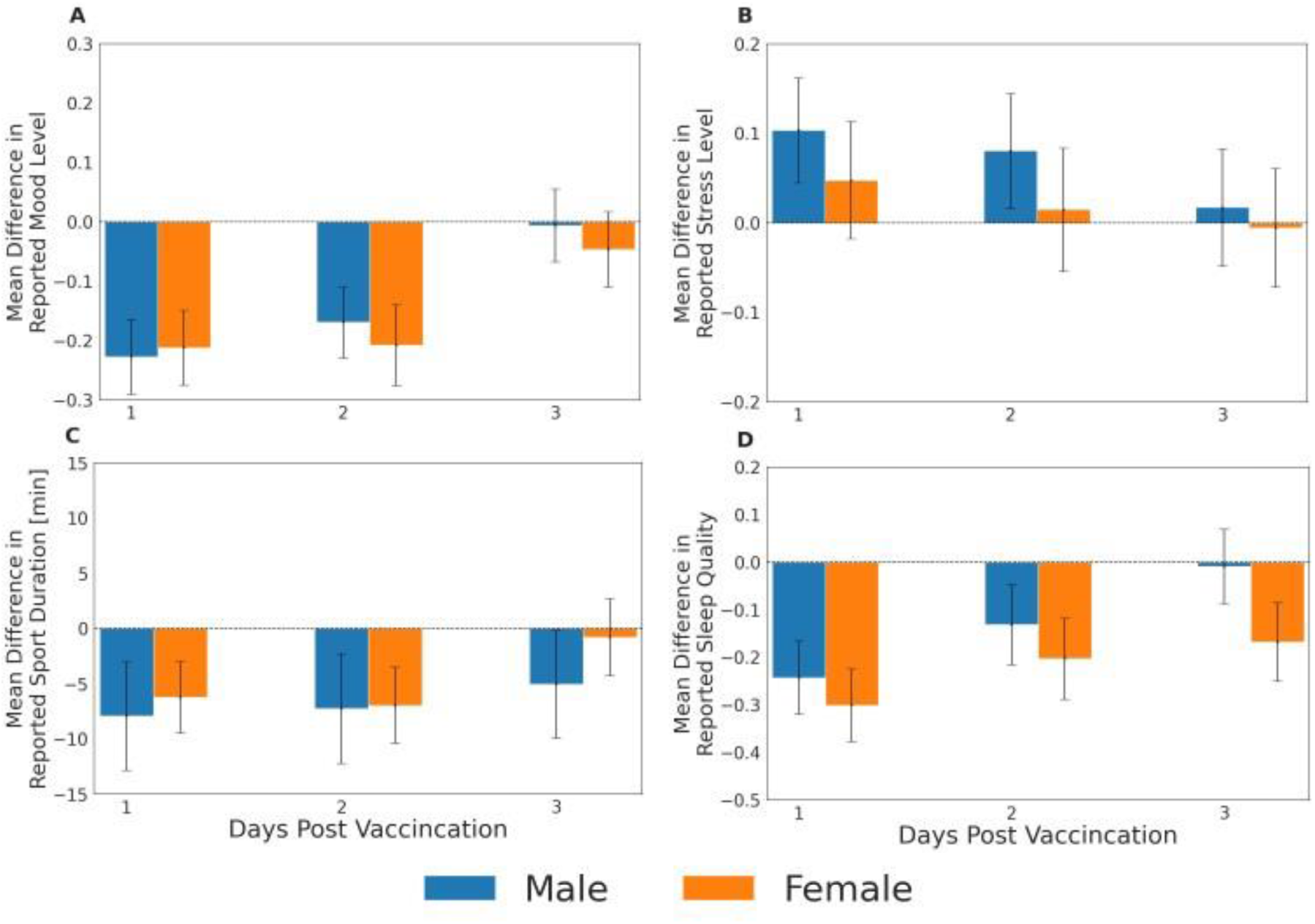
Changes in well-being indicators reported by participants through the mobile application stratified by gender. (A) mood level, measured on a 1-to-5 Likert scale. (B) Stress level, measured on a 1-to-5 Likert scale. (C) Sport duration, measured in minutes. (D) Sleep quality, measured on a 1-to-5 Likert scale. Changes in well-being indicators were calculated by subtracting the baseline values from the daily values. Error bars represent 90% confidence intervals. Horizontal dashed lines represent no change compared to baseline levels.

### Changes in reported well-being indicators – stratification by underlying medical condition

Changes in mood level and sleep quality observed during the first two days after the third vaccine dose were found to be higher for participants without underlying medical conditions compared to those with underlying medical condition (Figure S4).

**Figure S4.**
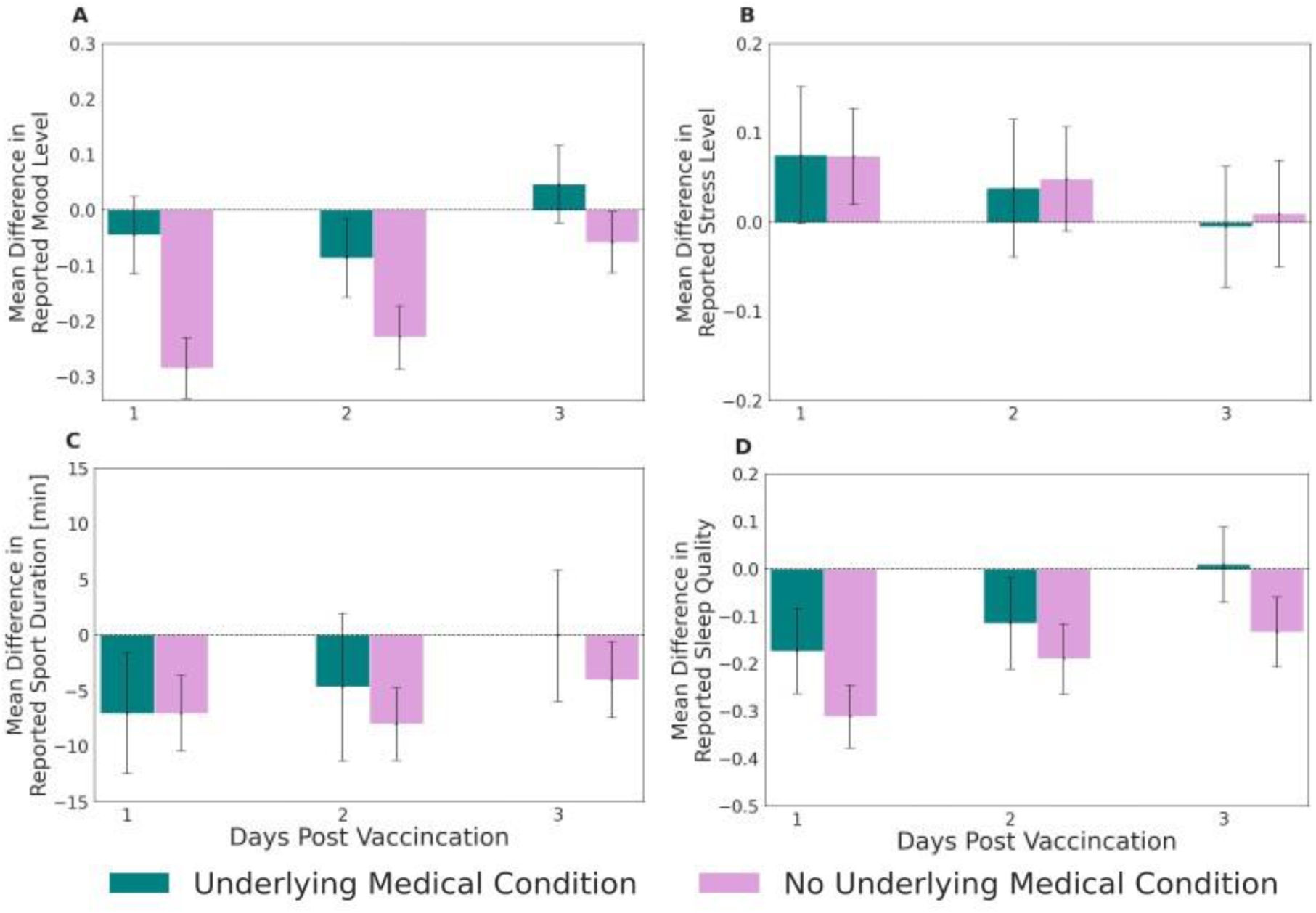
Changes in well-being indicators reported by participants through the mobile application stratified by underlying medical conditions. (A) mood level, measured on a 1-to-5 Likert scale. (B) Stress level, measured on a 1-to-5 Likert scale. (C) Sport duration, measured in minutes. (D) Sleep quality, measured on a 1-to-5 Likert scale. Changes in well-being indicators were calculated by subtracting the baseline values from the daily values. Error bars represent 90% confidence intervals. Horizontal dashed lines represent no change compared to baseline levels.

### Changes in physiological indicators – stratification by age group

Changes in physiological indicators after the third vaccine dose stratified by age group were consistent with those observed in the general population (considerable changes during the first two days after vaccine administration that faded nearly entirely after three days). These changes were found to be higher for participants younger than 50 years compared to those between 50 and 65 years, and consequently higher than those older than 65 years (Figure S5).

**Figure S5.**
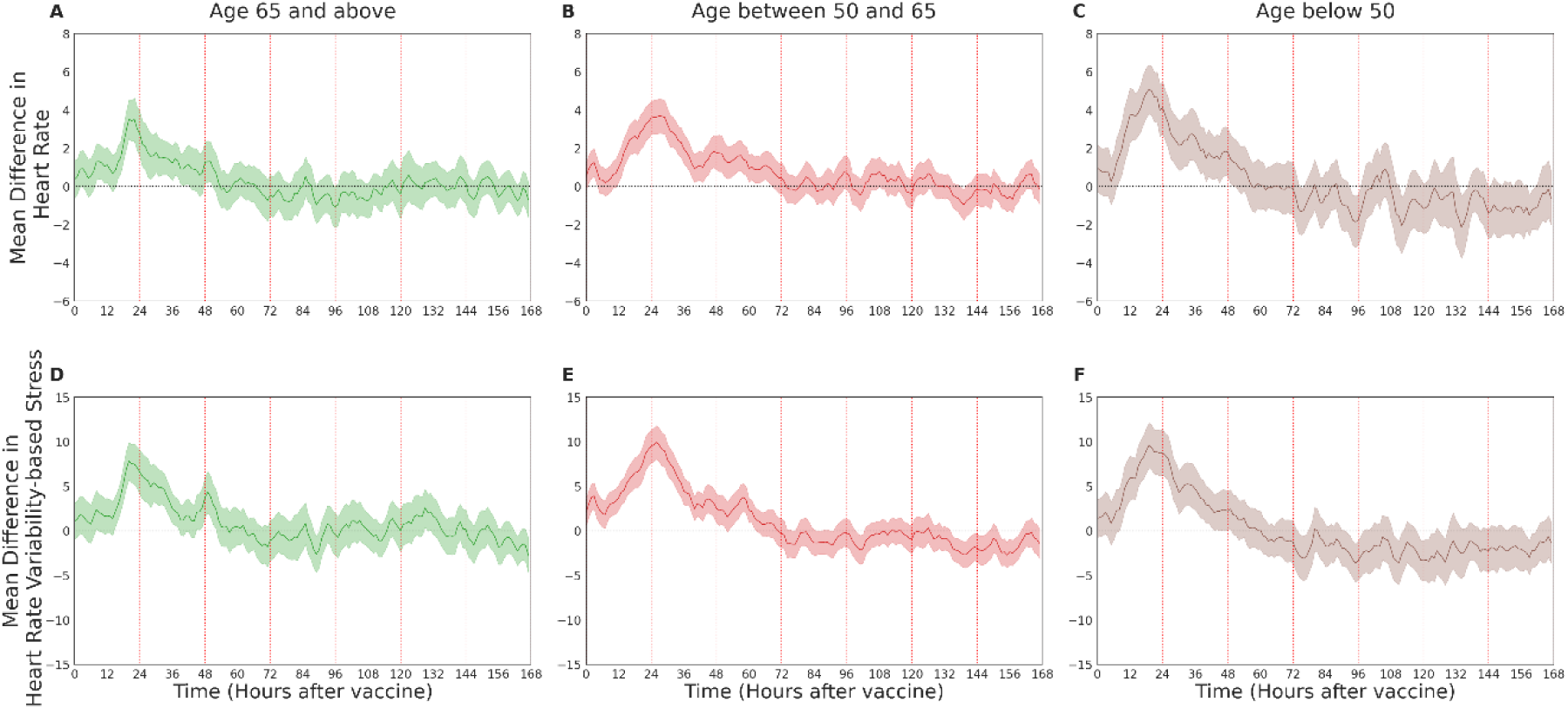
Changes in physiological indicators measured through the smartwatch stratified by age groups. Mean difference in heart rate and heart rate variability-based stress indicators following the third dose, recorded by a smartwatch, compared to their baseline levels: (**A** and **B**) heart rate, (**C** and **D**) heart rate variability-based stress. Mean values are depicted as solid lines; 90% confidence intervals are presented as shaded regions. The horizontal dashed line represents no change compared to the baseline levels, and vertical lines represent 24-hour periods.

### Changes in physiological indicators – stratification by gender

Changes in physiological indicators after the third vaccine dose stratified by gender were consistent with those observed in the general population (considerable changes during the first two days after vaccine administration that faded nearly entirely after three days). These changes were found to be higher for females compared to males (Figure S6).

**Figure S6.**
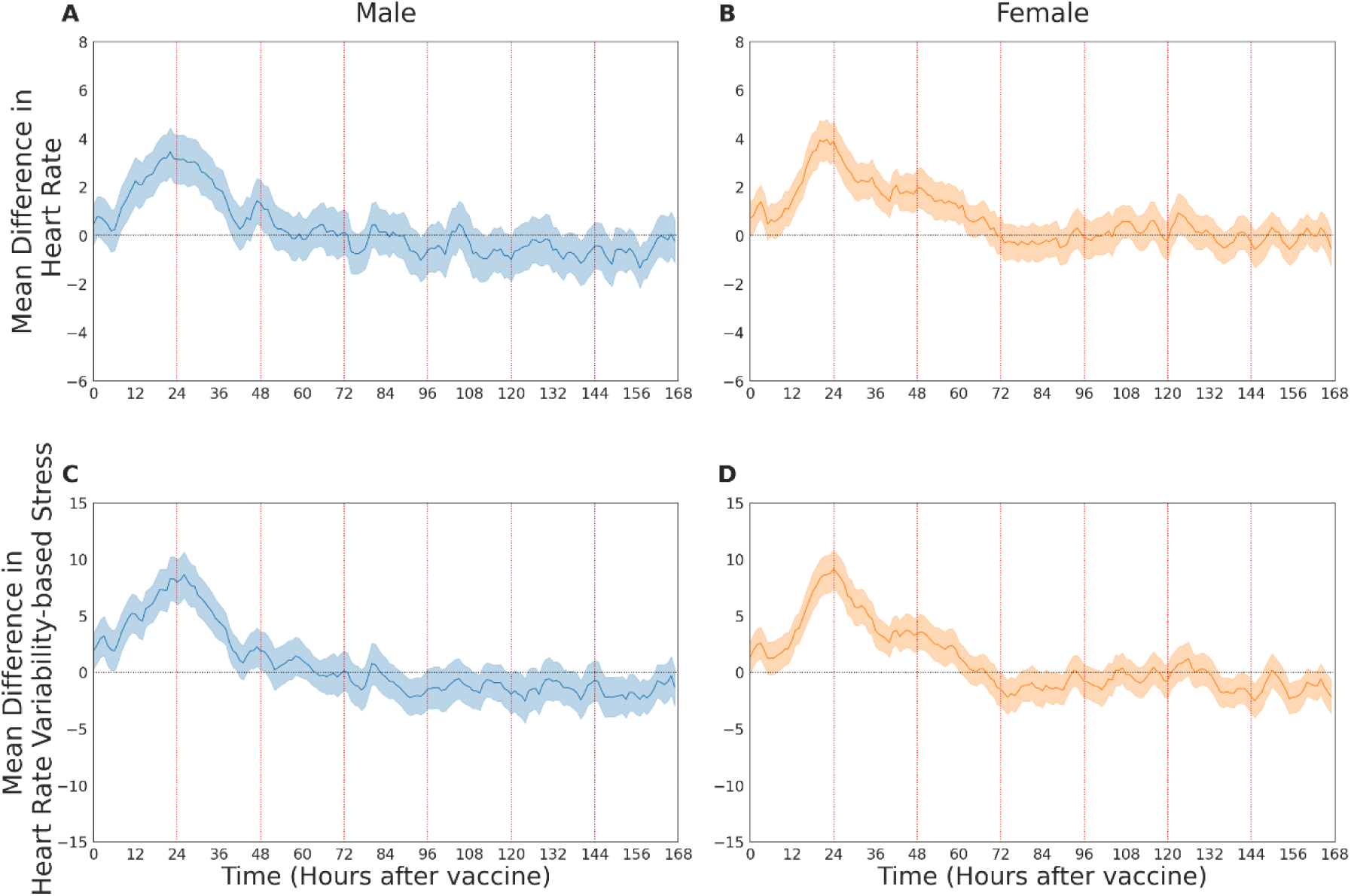
Changes in physiological indicators measured through the smartwatch stratified by gender. Mean difference in heart rate and heart rate variability-based stress indicators following the third dose, recorded by a smartwatch, compared to their baseline levels: (**A** and **B**) heart rate, (**C** and **D**) heart rate variability-based stress. Mean values are depicted as solid lines; 90% confidence intervals are presented as shaded regions. The horizontal dashed line represents no change compared to the baseline levels, and vertical lines represent 24-hour periods.

### Changes in physiological indicators – stratification by underlying medical condition

Changes in physiological indicators after the third vaccine dose stratified by underlying medical condition were consistent with those observed in the general population (considerable changes during the first two days after vaccine administration that faded nearly entirely after three days). These changes were found to be higher for participants without underlying medical conditions compared to those with underlying medical condition (Figure S7).

**Figure S7.**
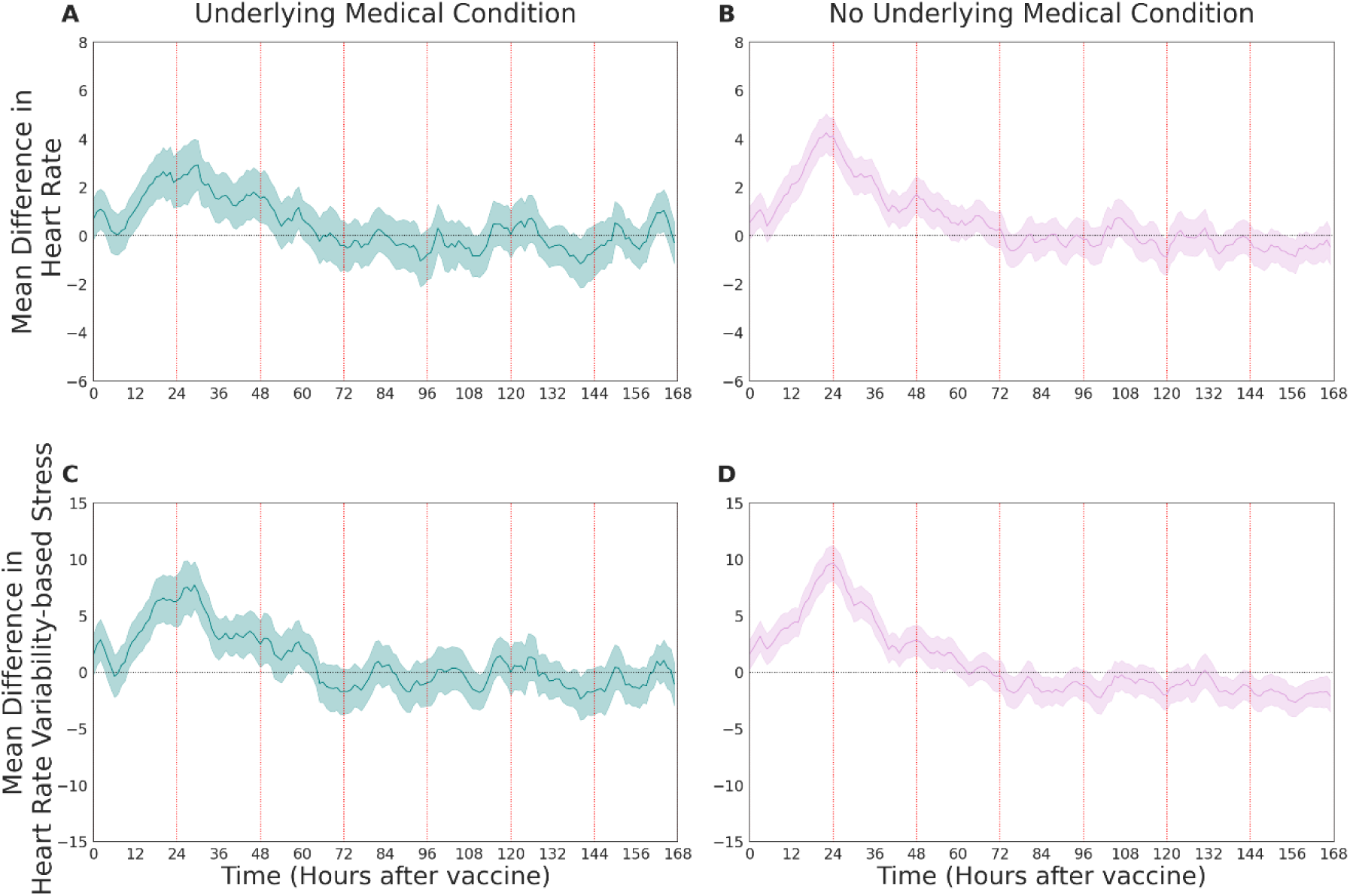
Changes in physiological indicators measured through the smartwatch stratified by underlying medical conditions. Mean difference in heart rate and heart rate variability-based stress indicators following the third dose, recorded by a smartwatch, compared to their baseline levels: (**A** and **B**) heart rate, (**C** and **D**) heart rate variability-based stress. Mean values are depicted as solid lines; 90% confidence intervals are presented as shaded regions. The horizontal dashed line represents no change compared to the baseline levels, and vertical lines represent 24-hour periods.

### 14 days analysis for self-reported local and systemic reactions after the third dose

We observe a sharp decline in reported local and systemic reactions following three days after the third vaccination dose, and nearly a complete halt within 14 days post-vaccination (Figure S8). Fatigue and headache were the most frequent reactions reported and lasted longer than the other reported reactions.

**Figure S8.**
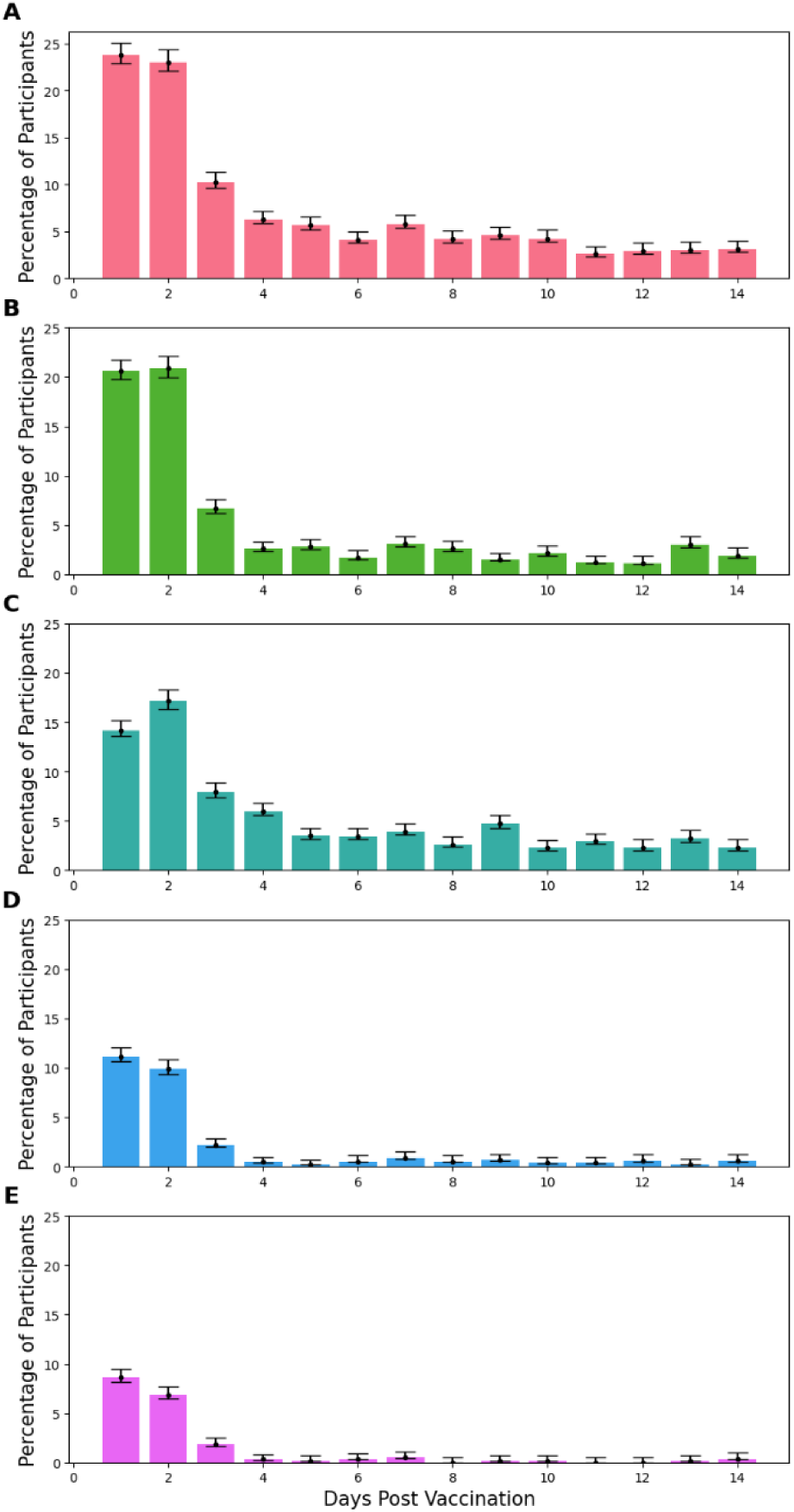
Most frequent local and systemic reactions reported by participants through the mobile application after the third dose. (**A**) fatigue, (**B**) muscle pain, (**C**) headache, (**D**) fever, and (**E**) chills. Error bars represent 90% confidence intervals.

### Number of participants receiving each vaccine dose

1,609 participants reported receipt of at least one of the three BNT162b2 mRNA COVID-19 vaccine shots after joining the study. Specifically, out of these 1,609 participants serving as the base cohort for our analyses, 223 received during the study period their first dose, 351 their second dose, and 1,344 their third dose (Table S1).

**Table S1.**
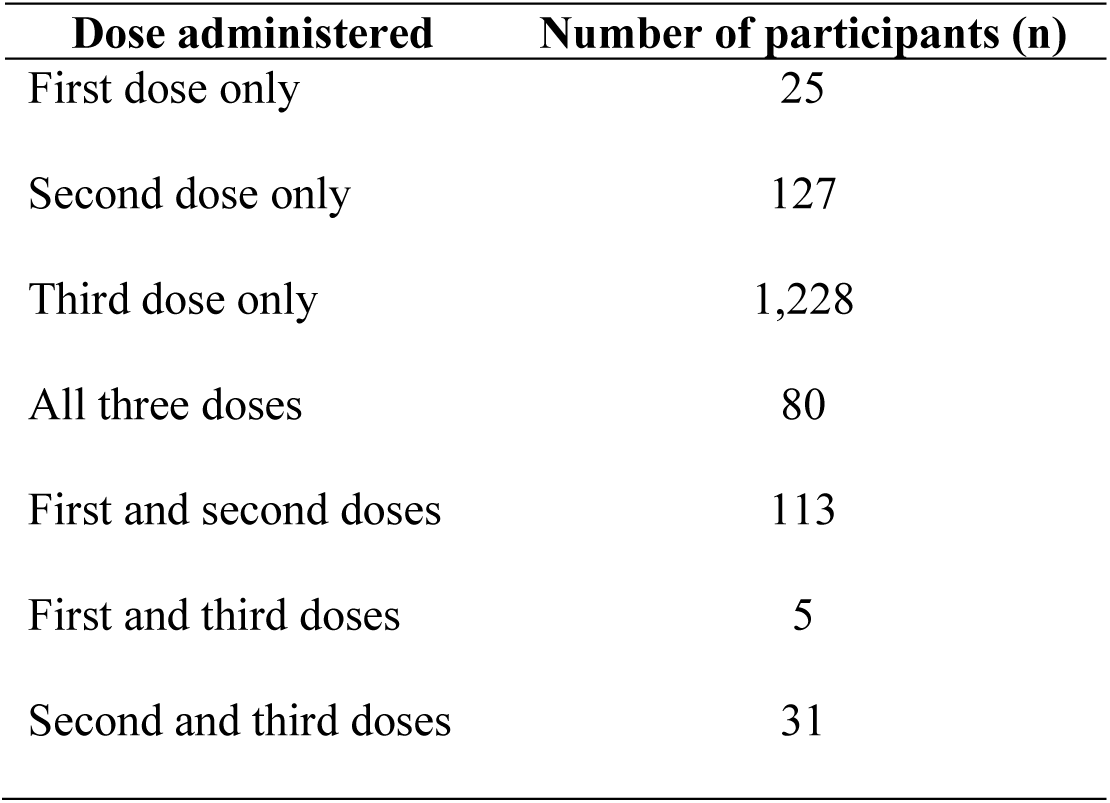
Number of participants receiving each vaccine dose

| **Dose administered** | **Number of participants (n)** |
| --- | --- |
| First dose only | 25 |
| Second dose only | 127 |
| Third dose only | 1,228 |
| All three doses | 80 |
| First and second doses | 113 |
| First and third doses | 5 |
| Second and third doses | 31 |

